# Local authority variation in uptake of the HPV vaccine in Greater Manchester and school-level factors: a cross-sectional ecological study

**DOI:** 10.1101/19013839

**Authors:** Steven L. Senior, Rebecca Fletcher, Paul Cleary, Siobhan Farmer

## Abstract

**Background:** Human papillomavirus (HPV) is the main cause of cervical cancer and contributes to a number of other cancers that affect both men and women. Vaccines exist that offer protection against the most common cancer-causing HPV types. In England, a school-based vaccination programme for girls has been in place since 2008 but vaccine coverage rates have declined since its introduction. Understanding variation between schools and between local authorities may help to inform quality improvement and guide policy development and commissioning.

**Methods:** Cross-sectional, ecological analysis of vaccine uptake among 164 schools representing 13,127 children in eight out of ten local authorities in Greater Manchester. Logistic mixed effects regression models were used to test for associations between school level factors and vaccine uptake, while allowing for variation between local authorities.

**Results:** In multivariable mixed effects models, lower vaccination rates were associated with: increasing numbers of children eligible for vaccination; increasing proportions of children eligible for free school meals; increasing proportions of children with English as an additional language; Ofsted ‘inadequate’ ratings; Christian and Muslim faith schools; independent schools and special schools.

**Conclusions:** Consistent with previous studies on vaccine uptake, this study identifies a number of factors that are associated with uptake of the school-based HPV vaccine programme. We also show that local authority variation remains after adjustment for the mix of schools in each area. This evidence may be used to guide vaccine providers, commissioners, and policymakers who want to increase uptake of the school-based HPV vaccine.

## INTRODUCTION

### Human Papillomavirus

Human papillomavirus (HPV) is a large family of over 300 double-stranded DNA viruses that infect the mucosa and skin (Bzhalava, Eklund, and Dillner 2015; McBride 2017). An estimated 30% of women will be infected with HPV within two years of becoming sexually active (Woodman et al. 2001). Point prevalance estimates in unvaccinated populations range from 35% to 45% (Howell-Jones et al. 2012; Baudu et al. 2014). Approximately 80% of people will be infected with HPV in their lifetime (Syrjanen et al. 1990; Chesson et al. 2014).

Most HPV infections clear within two years (Woodman et al. 2001; Muñoz et al. 2004). However prolonged infection is associated with warts and some types of cancer. At least 12 strains of HPV are known to cause cancer. HPV infections are responsible for an estimated 99% of cervical cancer, as well as being associated with penis, vulvovaginal, anal, and oropharyngeal cancers (McBride 2017).

### The HPV vaccine programme in England

Several vaccines are available that are effective in preventing HPV infections. These include bivalent (covering oncogenic HPV 16 and 18), quadrivalent (HPV 16 and 18 plus HPV 6 and 11 that cause genital warts) and nonavalent (covering HPV types 6, 11, 16, 18, 31, 33, 45, 52, and 58) vaccines.

The introduction of HPV vaccine programmes has been associated with marked falls in the prevalence of oncogenic HPV types, precancerous cervical lesions in girls, and genital warts in both sexes (Mesher et al. 2016; Palmer et al. 2019; Drolet et al. 2019).

In England, the HPV vaccine has been offered to girls in school year 8 and 9 (ages 12 - 14) since 2008. In 2019 the school HPV vaccine programme was expanded to include boys. The school-based programme uses a two-dose vaccination schedule, with a minimum six-month gap between the first and second dose. For children under 15 years old, two doses are considered necessary for full protection (Public Health England 2019a).

The target for uptake of the HPV vaccine in England is 80%. Uptake among girls was initially high at 90% nationally, but has since fallen to below 85% (Public Health England 2018). However, national figures mask variation in uptake between local authorities (local government areas in England). Within England uptake among girls in 2017/18 varied from 67.8% in Kensington and Chelsea to 95.3% in Rotherham (Public Health England 2019b).

Understanding the variation between local authorities is important for monitoring and improving performance. However, variation between local authorities may be explained by the make-up of the schools within their areas. Previous research has established a number of school-level factors that are associated with uptake (Fletcher et al. 2019; Tiley et al. 2019). These include the proportion of children eligible for free school meals (an indicator of poverty), the proportion of children for whom English is an additional language, school faith, the effectiveness of the school management (as measured by Ofsted, the schools inspectorate in England), and the type of school.

However, these previous studies have not considered the potential variation between local authorities, or the extent to which this variation is explained by variation in schools between areas. The aim of this study is to understand what factors are associated with variation in HPV vaccine uptake between schools in Greater Manchester, and whether the variation between local authorities in uptake of the HPV vaccine in Greater Manchester is explained by variation in school level factors, and to replicate previous results using local data.

## METHODS

### Study design

This is an ecological, cross-sectional study using routinely available data.

### Setting

Greater Manchester is a city region in the North West of England. In 2018 it had a population of approximately 2.8 million (Office of National Statistics 2019). It is made up of ten local authorities. The population is more deprived than average for England (Greater Manchester Health and Social Care Partnership 2017; Public Health England 2019b).

### Data Sources and variables

School level data on vaccine uptake was provided by eight out of ten local authorities in Greater Manchester. This data was extracted from local Child Health Information Systems. Data was not available for Salford and Wigan local authorities. Schools where data on key variables was missing were excluded from analysis.

Data on school characteristics was downloaded from the Department for Education’s website (Department for Education 2019). Data on school performance was downloaded from the Ofsted website (Ofsted 2019). Where data was missing in the downloaded datasets, we checked Government and Ofsted websites and manually added this data if it was available. Table 1 summarises the variables used in this study.

**Table 1.** Description of independent variables used.

| **Variable** | **Description** |
| --- | --- |
| School size | The total number of pupils attending the school. |
| Percentage of pupils eligible for free school meals | The percentage of the total school population that are eligible for free school meals. This is used as a proxy for poverty. |
| Percentage of pupils with English as an additional language | The percentage of the total school population where English is not the first language, often meaning that English is not the language spoken by the child's parents. |
| Percentage receiving support for special educational needs | The percentage of pupils receiving support for special educational needs. |
| Ofsted rating | The overall effectiveness rating reported at the school's most recent Ofsted inspection. Levels: 'outstanding', 'adequate', 'requires improvement', and 'inadequate'. The reference for analysis here is 'outstanding'. |
| School faith | The school's religious affiliation. The reference for analysis is 'no faith'. |
| Type of school | The type of school. Levels: Community schools (local authority employs the staff, owns the buildings, and sets admissions policies), Foundation and voluntary schools (local authority funds the school, but a governing body employs the staff and sets the admissions policy), Academy schools and free schools (funded by government and independent of local authority). |

Ofsted is the Office for Standards in Education. It inspects organisations providing education services in England.

### Statistical Analysis

Vaccination rates were calculated as the number of children receiving two doses of HPV vaccine divided by the number of children eligible for vaccination in the school. This included vaccinated children who received the vaccine through their GP rather than in school.

We linked uptake data with DFE and Ofsted data using school unique reference numbers. Where unique reference numbers were not available, data was matched on school names.

We used univariable and multivariable mixed-effects logistic regression models (schools nested within local authorities) to test for associations between school level variables and HPV vaccine uptake. Local authority random intercepts were included in the multivariable model to understand residual variation between local authorities after accounting for school-level variation and to adjust for non-independence of error terms (as schools within the same local authority are likely to be more similar than those more geographically distant). Independent variables were centred and scaled to mean 0 and standard deviation 1 to aid model fitting. This means that the odds ratios reported reflect the change in the odds of vaccination for every one standard deviation change in the independent variable.

Previous studies have found an interaction between the proportion of children eligible for free school meals (an indicator of levels of poverty and deprivation among children at the school) and the proportion of children with English as a second language. We include this interaction term here for comparability with other studies.

Statistical analysis was conducted in R version 3.5.3 (R Core Team 2019). The tidyverse and readxl packages were used for data manipulation (Wickham 2017; Wickham and Bryan 2019) and the lme4 package was used for fitting mixed-effects logistic regression models (Bates et al. 2015). Logistic regression models were fitted to the proportion of eligible children receiving two doses of the HPV vaccine by using the number of eligible children (the denominator) as weights in the regression model.

### Ethical approval

Ethical approval was not necessary for this study because it was conducted using only aggregated data and is a service evaluation.

## RESULTS

There were 174 schools in total, comprising 13,159 eligible pupils. 10 schools were excluded because there was missing data for key variables. This left 164 schools that were included in the study, comprising 13,127 pupils.

The overall vaccination rate in schools across the eight local authority areas included in the study was 81.1% (range 0% to 100%, interquartile range 71.1% to 89.6%).

The vaccination rate for local authorities ranged from 67.2% to 89.8%. Table 2 shows the characteristics of the schools included in this study.

**Table 2.** Characteristics of schools included in the study, Greater Manchester, 2017/18.

| **Variable** | **Median** | **IQR** |
| --- | --- | --- |
| Number of eligible pupils per school | 81 | 72.5 |
| Total number of pupils at school | 863.5 | 741.5 |
| % eligible for free school meals | 17.6 | 18.6 |
| % with English as an additional language | 10.1 | 26.3 |
| % receiving support for special educational needs | 7.8 | 9.4 |
| **Variable** | **N** | **%** |
| Number of schools | 164 | 100 |
| Ofsted rating |  |  |
| Outstanding | 36 | 21.95 |
| Good | 73 | 44.51 |
| Requires improvement | 28 | 17.07 |
| Inadequate | 14 | 8.54 |
| Not available | 13 | 7.93 |
| Faith |  |  |
| No faith | 115 | 70.12 |
| Christian | 39 | 23.78 |
| Jewish | 1 | 0.61 |
| Muslim | 9 | 5.49 |
| School type |  |  |
| Community school | 27 | 16.46 |
| Academy or free school | 67 | 40.85 |
| Independent school | 23 | 14.02 |
| Special school | 18 | 10.98 |
| Voluntary or foundation school | 29 | 17.68 |

IQR = interquartile range.

Table 3 shows the unadjusted odds ratios and 95% confidence intervals from the univariable logistic regression model.

**Table 3.** Unadjusted odds ratios and 95% confidence intervals for school characteristics and HPV vaccine uptake, Greater Manchester, 2017/18

| Variable | Odds ratio | 95% Conf. Int. | P-value |
| --- | --- | --- | --- |
| Number of eligible pupils | 1.000 | 1 to 1 | 0.425 |
| % eligible for free school meals | 0.841 | 0.79 to 0.9 | < 0.001 |
| % pupils with English as an additional language | 0.866 | 0.83 to 0.9 | < 0.001 |
| % receiving support for special educational needs | 1.007 | 0.95 to 1.07 | 0.826 |
| Ofsted 'Outstanding' | 1.000 | - | - |
| Ofsted 'good' | 1.093 | 0.98 to 1.22 | 0.12 |
| Ofsted 'requires improvement' | 1.086 | 0.95 to 1.24 | 0.235 |
| Ofsted 'inadequate' | 1.084 | 0.91 to 1.28 | 0.353 |
| Not available | 1.286 | 1.01 to 1.64 | 0.04 |
| Non-faith school | 1.000 | - | - |
| Christian faith school | 1.028 | 0.93 to 1.14 | 0.588 |
| Jewish faith school | 6.827 | 1.67 to 27.85 | 0.007 |
| Muslim faith school | 0.625 | 0.48 to 0.81 | 0.001 |
| Community school | 1.000 | - | - |
| Academy or free school | 0.800 | 0.71 to 0.9 | < 0.001 |
| Independent school | 0.896 | 0.73 to 1.1 | 0.288 |
| Special school | 0.191 | 0.13 to 0.28 | < 0.001 |
| Foundation or voluntary school | 1.112 | 0.96 to 1.28 | 0.147 |

Odds ratios for Ofsted ratings are relative to schools with an 'outstanding' rating. Odds ratios for faiths are relative to schools with no faith affiliation. Odds ratios for school types are relative to community schools.

In unadjusted analyses, Jewish faith was associated with higher odds of vaccination among pupils (although it is important to note that there was only one Jewish faith school in the data set). The proportion of children eligible for free school meals, the proportion of children for whom English is an additional language, academy and free schools, and special schools were associated with lower odds of vaccination among pupils. Vaccination uptake did not vary significantly by Ofsted rating, but schools for which Ofsted ratings were not available (independent schools that are inspected by the Independent Schools Inspectorate) appeared to have higher odds of vaccination among their pupils.

In multivariable analyses, a model with local authority random effects performed significantly better than a single-level model without local authority random effects (Chi squared = 366.6 on 1 degree of freedom, p < 0.001). In a model containing only local authority random intercepts and no school-level explanatory variables, the standard deviation of the random intercepts was 0.445. Adding the school level variables increased the standard deviation of the random intercepts to 0.503.

Together, this suggests that most variation between local authorities in HPV vaccine uptake that is not explained by the mix of the schools within their areas based on the school characteristics that we have used in this study.

Table 4 shows the adjusted odds ratios and 95% confidence intervals from the multivariable mixed effects model.

**Table 4.**
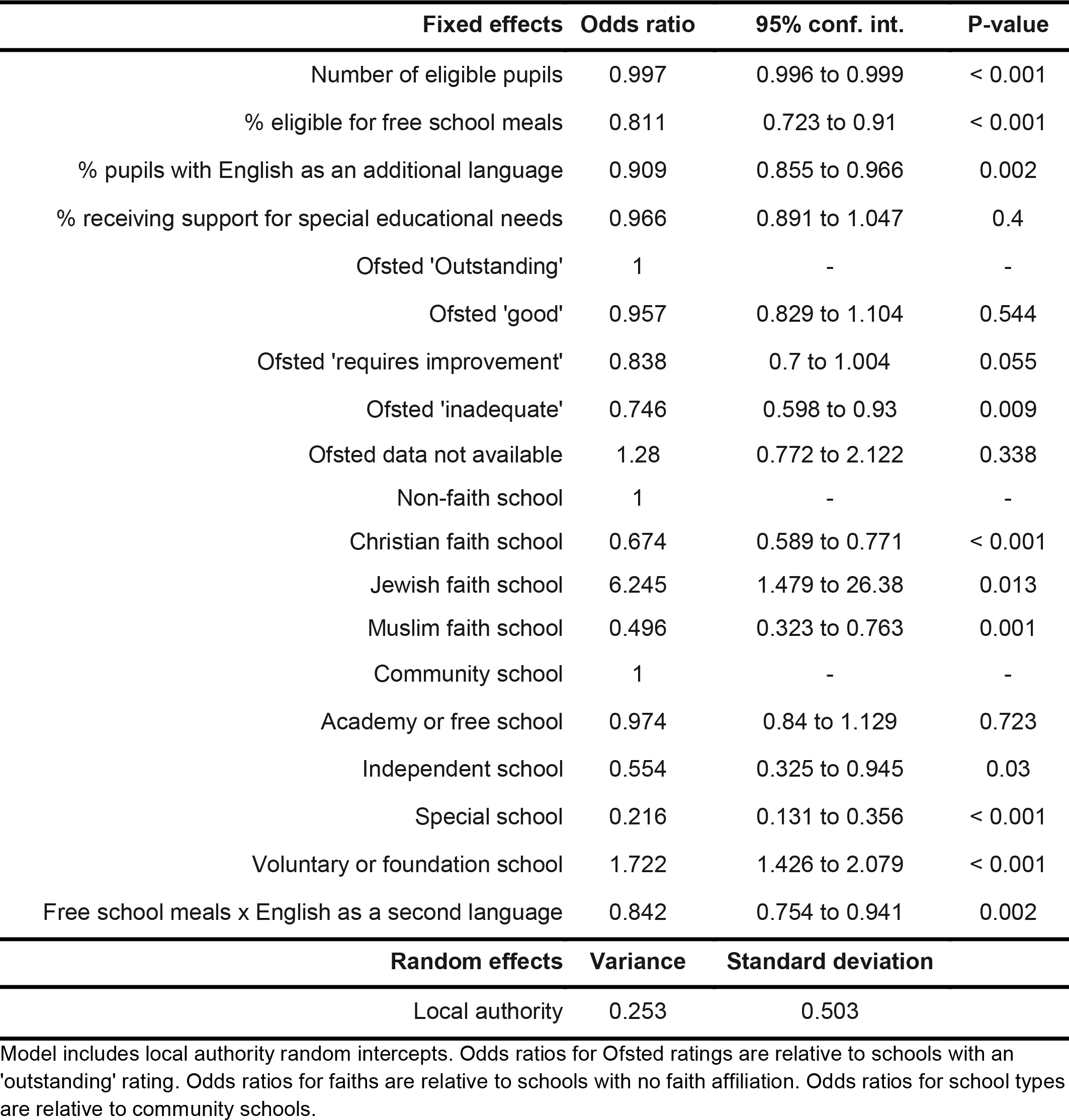
Adjusted odds ratios and 95% confidence intervals for school characteristics and HPV vaccine uptake, Greater Manchester, 2017/18.

In the multivariable random intercepts model, Jewish faith and voluntary or foundation school status was associated with higher odds of vaccination. Lower odds of vaccination were associated with: increasing numbers of children eligible for vaccination, a higher proportion of children eligible for free school meals, a higher proportion of children for whom English is an additional language, Ofsted ‘inadequate’ ratings, Christian faith, and independent or special school status. In addition, while ‘inadequate’ was the only category of Ofsted rating that was significantly associated with lower odds of vaccination among pupils, there appears to be a trend towards decreasing odds of vaccination across the other Ofsted ratings.

There was a significant interaction between the proportion of children eligible for free school meals and the proportion for whom English was an additional language. This interaction was negative, indicating that its effect was to exacerbate an already negative association between those two factors and HPV vaccine coverage. The total effect of a one standard deviation increase in both the proportion eligible for free school meals and the proportion for whom English is an additional language was an odds ratio of 0.621.

## DISCUSSION

### Vaccine uptake

Overall, the HPV vaccine coverage rate remains above the national target of 80%, although the falls in coverage seen in previous years appear to have continued.

### Variations in HPV vaccine uptake between schools

This study identifies a number of school-level characteristics that appear to explain some of the variation in HPV vaccine uptake between schools.

Consistent with other studies on school-based vaccine uptake, the proportion of children in a school who are eligible for free school meals and the proportion for whom English is an additional language were associated with lower vaccine uptake (Fletcher et al. 2019). In addition, we found a significant interaction between these two variables, such that in schools with both a high proportion of children eligible for free school meals and with English as an additional language, HPV vaccine coverage was lower still. This differential uptake is likely to result in inequalities in cancer rates between more and less affluent children and children of recent migrants. As earlier studies have pointed out, the effect of English as an additional language may be explained by those children’s parents having English as an additional language and this being a barrier to communication and parental consent. Previous studies have found that more knowledge about HPV was associated with greater intent to vaccinate among parents in Northern England (Sherman and Nailer 2018). This suggests that translation of vaccination information leaflets may be an appropriate strategy to increase parental consent for vaccination among families whose first language is not English. Alternatively, in both more deprived families and families where English is a second language, attitudes to vaccination may be different than in more affluent and English speaking families. One systematic review of barriers to HPV vaccination found that parental beliefs about the effect of vaccinating against a sexually transmitted infection on sexual behaviour may also affect parental consent (Holman et al. 2014).

The lower vaccination rates observed in Christian and Muslim faith schools may also reflect attitudes to vaccination against what is primarily a sexually-transmitted infection. This finding is consistent with other studies (Fletcher et al. 2019; Tiley et al. 2019). However, the international evidence base on the effect of religion on attitudes to HPV vaccination is mixed, with some studies finding that religious beliefs are associated with higher vaccination rates (Shelton et al. 2013), other studies finding no association (Brabin et al. 2006; Lenselink et al. 2008), and others still finding religious beliefs were associated with lower vaccination rates (Marlow, Waller, and Wardle 2007; Marlow et al. 2009). UK-based studies tend to find a negative association between religious faith and HPV vaccine acceptability, and qualitative studies suggest that conservative social attitudes may play a role (Marlow, Waller, and Wardle 2007; Marlow et al. 2009; Laura et al. 2009; Gordon, Waller, and Marlow 2011).

The higher vaccination rates associated with Jewish schools observed here are at odds with the findings of a broader study (Tiley et al. 2019), but there was only one Jewish school in this dataset. This suggests that there may be important variation within categories of faith schools, and it may be possible to work with faith schools with high levels of uptake to understand how best to improve vaccine uptake among other schools that share the same faith.

Another difference between this study and other studies of school-based vaccine uptake was the observation that increasing numbers of pupils eligible for vaccination was associated with lower vaccination rates. It is worth noting that the effect, while statistically significant, was small (a 1 standard deviation increase in the number of eligible pupils was associated with a 0.3% relative decrease in the odds of vaccination among eligible pupils).

Consistent with previous research, we found that schools with the worst overall effectiveness rating (‘inadequate’) had lower vaccine uptake rates (Fletcher et al. 2019). While the effect was statistically significant only for the lowest Ofsted rating, the pattern was of decreasing odds of vaccination with decreasing overall school effectiveness ratings. The lack of a significant association between the ‘good’ and ‘requires improvement’ Ofsted ratings and HPV vaccine uptake may therefore reflect a type II error, and a larger study may be needed to understand whether there is an overall trend for lower uptake to be associated with worse Ofsted effectiveness ratings.

The variables identified here are unlikely to explain the fall in the uptake of the HPV vaccine. To the extent that data is available, there is little evidence that these variables have changed over time in a way that would explain the fall in vaccination rates. However, it is possible that the effect of these variables over time has changed. For example, while the proportion of children who are eligible for free school meals across Greater Manchester has decreased since 2014 (Public Health England 2019b), it is possible that schools with more children eligible for free school meals have seen bigger falls in coverage. Further analysis using longitudinal data would be needed to test this hypothesis.

### Variations in HPV vaccine uptake between local authorities

As local authorities are responsible for commissioning school nursing and other early years health services, a better understanding of variation between local authorities may help to guide local and national policy and commissioning.

Despite meeting the national target overall, three of the eight local authorities studied here were below the 80% target. The results presented here suggest that this variation is not explained by variation in the make-up of schools within each area according to the school characteristics included in this study.

### Strengths of this study

This study is the first to our knowledge to examine school-level factors that affect variation in HPV vaccine uptake while allowing for variation between local authorities in uptake. This approach can help to understand variation in vaccination between local authorities, while adjusting for the make-up of schools in each area. This approach can support better assessment of the performance of HPV vaccine delivery within each local authority area. The use of mixed models also helps to adjust for the fact that HPV vaccine uptake rates in schools in the same local authority area may not be independent.

We also consider a broader set of school-level characteristics than previous studies (Fletcher et al. 2019; Tiley et al. 2019). In particular, where Tiley et al use area-level deprivation scores, we use the proportion of children who are eligible for free school meals within each school’s student population. This may better reflect the level of poverty among the children who attend the school.

### Limitations of this study

This is an ecological study and as such, the results here should be treated with caution. Further research using individual level data would allow for more confident inferences about the role of pupil and family factors on vaccine uptake. This would complement qualitative and survey evidence on factors affecting HPV vaccine consent (Sherman and Nailer 2018; Laura et al. 2009; Gordon, Waller, and Marlow 2011).

The study is also limited in its sample of local authority areas. Data was only available for eight of the ten local authorities in Greater Manchester. However the results are broadly consistent with a study that included 42 out of 152 upper-tier local authorities in England (Tiley et al. 2019).

There are other school-level factors that were not included in this study that may also influence vaccine uptake. For example, we did not include data here on school expenditure per pupil, as this was not available for all the schools in the data set.

This study also suffers from a lack of local authority-level explanatory variables. The inclusion of such data may help to explain the apparent variation between local authorities that is not explained by school or individual and family factors. Such data might include configuration of and spending on vaccine providers, school nurses, and other childrens health services.

As the school-based HPV vaccine programme in England has now been running for over ten years, longitudinal studies of HPV vaccine uptake are possible. The use of longitudinal datasets may allow for a better understanding of the school and local authority level factors that affect HPV vaccine uptake among communities in England.

## Data Availability

School-level HPV uptake data were extracted from local child health information systems and are not publicly available. School-level factors were downloaded from the Department for Education and the Ofsted websites.

## Acknowledgements

We are grateful for the support of the local authorities who provided the HPV vaccination data that were used in this study, and to David Holderness of NHS England (Greater Manchester) for support with data extraction.

